# Development and evaluation of machine learning models for classifying heart failure using clinical and demographic data in Sidama, Ethiopia

**DOI:** 10.64898/2026.11.26.26369031

**Authors:** Melaku Haile Likka, Sura Markos, Solomon Mamo Banteywalu, Taye Gari

## Abstract

**Background and objective:** Heart failure (HF) is clinically heterogeneous and challenging to diagnose, particularly in settings with limited access to specialized investigations. Machine-learning (ML) models using routinely available clinical data may improve HF identification, although evidence from sub-Saharan Africa is limited. This study developed and evaluated ML models to classify HF among patients attending four public hospitals in Sidama Region, Ethiopia

**Methods:** A cross-sectional ML study was conducted using data collected from January–March 2025 in four public hospitals in Sidama region. The analytic dataset included 716 participants: 386 with a documented HF diagnosis recruited from cardiac follow-up clinics and 330 without a documented HF diagnosis recruited from outpatient departments (OPD). Forty candidate predictors were considered. Seven classifiers were compared using stratified cross-validation in the training dataset after an 80:20 stratified split. Preprocessing and ReliefF feature selection were performed within the model-development folds. The leading three models underwent randomized hyperparameter optimization. The final model was selected using cross-validated ROC-AUC and evaluated on the held-out test set. Discrimination, threshold-specific performance, calibration, decision-curve analysis and feature importance were assessed.

**Results:** Extra Trees achieved the highest cross-validated ROC-AUC (0.955) and test-set ROC-AUC (0.954; 95% CI: 0.918–0.982) and PR-AUC (0.966; 95% CI: 0.938–0.987). On the test set, accuracy was 90.3% (95% CI: 85.4%–94.4%), sensitivity 87.2% (79.3%–94.1%), specificity 93.9% (87.5%–98.6%), precision 94.4% and negative predictive value 86.1%. The Brier score was 0.082; the calibration intercept was 0.025 and slope 1.352. Prior history of heart attack or stroke was the most influential predictor in permutation-importance and SHAP analyses. Logistic regression had slightly higher test-set accuracy (91.0%) than Extra Trees.

**Conclusion:** Extra Trees demonstrated high internal discrimination of documented HF status in this selected hospital-based sample. Routinely obtainable demographic and clinical characteristics may help distinguish patients with established HF from other OPD attendees. However, external validation, reliable outcome ascertainment, assessment of post-diagnostic predictors, and recalibration are necessary before clinical application.

## Introduction

Heart failure (HF) is a clinical syndrome characterized by current or previous symptoms and signs resulting from cardiac structural or functional abnormalities, supported by objective evidence of cardiac congestion or elevated natriuretic peptide levels [1]. It represents a major clinical manifestation of cardiovascular diseases (CVD), arising from diverse underlying cardiac conditions, particularly ischemic heart disease and hypertensive heart disease, which contribute substantially to its global burden [2]. Globally, CVD account for approximately 19.8 million deaths in 2022, equivalent to 32% of all deaths worldwide, with more than three-quarters occurring in low-and middle-income countries (LMIC) [3]. In 2021, an estimated 55.5 million people were living with HF, representing a 118.2% increase in prevalent cases since 1990, with its global prevalence and disability burden projected to rise through 2050 [2].

The clinical presentations of HF vary according to the underlying cardiac pathology, disease severity, and degree of cardiac congestion. Common symptoms include dyspnea, orthopnea, paroxysmal nocturnal dyspnea, fatigue, reduced exercise tolerance, and peripheral swelling, while clinical signs include elevated jugular venous pressure, pulmonary crackles, peripheral edema, tachycardia, and a third heart sound [1,4]. Important risk factors include advanced age, hypertension, ischemic heart disease, previous myocardial infarction, diabetes mellitus, chronic kidney disease, and atrial fibrillation. Obesity, excessive alcohol consumption, valvular heart disease, and cardiomyopathies are additional contributing factors [1,4,5]. The diversity of clinical presentations and associated risk factors complicates HF assessment.

Despite its substantial clinical and public health burden, HF recognition and diagnosis remain challenging because of its heterogeneous presentation and overlapping symptoms with other cardiovascular and non-cardiovascular conditions. Dyspnea, fatigue, reduced exercise tolerance, and peripheral edema are not specific to HF and may occur in patients with pulmonary diseases, renal dysfunction, anemia, and other clinical conditions [4,6]. Although clinical history and physical examination are essential initial components of the diagnostic assessment, individual symptoms and signs have limited diagnostic accuracy when considered in isolation [4,6].

Consequently, contemporary diagnostic guidelines recommend combining clinical assessment with electrocardiography, circulating natriuretic peptide measurements, and cardiac imaging, particularly transthoracic echocardiography, to establish cardiac structural and functional abnormalities [4,5].

In sub-Saharan Africa (SSA), HF is a major CVD and exhibits a distinct epidemiological pattern compared with high-income regions. Patients in SSA tend to develop HF at relatively younger ages, and its underlying causes include hypertensive heart disease, cardiomyopathies, rheumatic heart disease, and ischemic heart disease [7–9].

In Ethiopia, hospital-based studies have reported a diverse spectrum of HF etiologies, including dilated cardiomyopathy, coronary artery disease, hypertensive heart disease, rheumatic heart disease, ischemic heart disease, and cor pulmonale [10,11]. However, the hospital-based nature of existing evidence limits understanding of the broader population burden and clinical spectrum of HF. These limitations are compounded by gaps in cardiovascular service readiness and access to specialized diagnostic services [8,12]. Consequently, identifying patients with HF and distinguishing them from patients presenting with other clinical conditions can be particularly challenging, highlighting the need for improved approaches to HF identification in resource-constrained healthcare settings.

Machine learning (ML), a branch of artificial intelligence (AI), offers opportunities to improve clinical assessment by identifying complex patterns and relationships among multiple patient characteristics that may not be readily apparent through conventional analytical approaches [13,14]. Machine learning algorithms can integrate demographic, clinical, and laboratory information to support the identification and classification of individuals with specific clinical conditions, including HF [15,16]. Previous studies have demonstrated the potential of ML and AI for HF identification, classification, and outcome prediction using electronic health records (EHRs), echocardiographic findings, and biomarkers [17–19]. In SSA, ML has been applied to predict HF-related hospital readmission in Rwanda, while AI-enabled digital auscultation has been investigated for detecting HF with reduced ejection fraction [20,21].

Despite these advances, the application of ML for HF identification in resource-constrained healthcare settings is underexplored. Although an Ethiopian study has demonstrated the application of ML to CVD prediction using routinely collected hospital data, it focused on broader CVD classification rather than HF specifically [22]. Evidence on the ability of routinely available demographic and clinical characteristics to discriminate between patients with established HF and those without the condition is limited, particularly in SSA and Ethiopia. Evaluating such approaches may provide useful evidence on their potential to support HF identification where access to specialized diagnostic investigations is variable.

Therefore, this study aimed to develop and evaluate ML models for classifying patients with and without established HF using demographic and clinical characteristics among patients attending public hospitals in Sidama Region, Ethiopia. Specifically, it compared the classification performance of different ML algorithms and identified the demographic and clinical characteristics contributing most to the discrimination of HF status.

## Methods

### Study design and prediction objective

A ML prediction study was conducted using cross-sectional data collected from January–March 2025 in four public hospitals with cardiac clinics in Sidama region, Ethiopia: Hawassa University Comprehensive Specialized Hospital and Adare General Hospital in Hawassa City Administration, Leku General Hospital in Shebedino District, and Yirgalem General Hospital in Dale District.

Sidama is one of Ethiopia’s 12 regional states, with Hawassa as its capital. The region had an estimated population of 4.8 million in 2024 [23,24] and 22 public hospitals at the time of the study: one comprehensive specialized, seven general, and 14 primary hospitals.

The study aimed to classify participants by documented HF status using routinely collected demographic, clinical, medication-related, and behavioral characteristics.

### Study population and participant recruitment

The source population comprised patients attending the cardiac clinics and OPD of the participating hospitals during the study period. The study population included patients aged ≥15 years receiving HF follow-up care or attending OPD.

The target variable was HF status, coded as a binary outcome (heartFDx: 1 = HF; 0 = no HF). Participants with HF were recruited from cardiac clinics on designated follow-up days based on a pre-existing clinical diagnosis documented in their medical records and patient registers. These patients were attending the clinics for follow-up, preventive, and other ongoing care. Participants without documented HF were recruited from OPD and were classified as non-HF if their medical records contained no documented HF diagnosis and they were attending for another infectious or noninfectious condition. Accordingly, HF status reflected pre-existing clinical documentation at recruitment, with no new diagnostic assessment or outcome adjudication undertaken for this study. No formal blinding of outcome ascertainment to candidate predictor information was implemented.

Candidate predictors included demographic, clinical, medication-related, and behavioral characteristics.

### Data collection and pre-testing

The data-collection instrument was pretested on 40 individuals at Motite Fura Primary Hospital to assess its clarity, comprehensibility, and sequence. Necessary modifications were made before the main study, and pretest participants were excluded from the main study.

### Data preparation

The final analytic dataset comprised 716 observations and 41 variables, including the binary target variable (heartFDx: 0 = no HF; 1 = HF) and 40 candidate predictors. Predictors included numerical and categorical variables, with categorical variables numerically encoded as required by the classification algorithms.

Data completeness, consistency, and outcome distribution were assessed before model development. Structural missingness in conditional follow-up variables was addressed through logical coding based on preceding responses. For example, participants reporting no moderate physical activity were assigned “No moderate exercise” for the corresponding frequency variable, while those who had never consumed alcohol were assigned “None” for the type of alcoholic beverages consumed. No other missing observations were identified. Following logical coding, the final analytic dataset contained no missing values; therefore, statistical imputation was unnecessary. The dataset included 330 participants without HF (46.1%) and 386 with HF (53.9%). Given the relatively balanced outcome classes, no dataset-wide class-imbalance correction or synthetic minority oversampling technique (SMOTE) was applied, although classifiers incorporating algorithm-specific class weighting or resampling were retained.

Feature selection was performed using ReliefF within the model-development pipeline rather than preliminary bivariate significance testing. This ensured that feature selection used only data available within each development fold, minimizing information leakage during cross-validation.

### Data partitioning

The dataset was divided into training (80%) and independent test (20%) sets using stratified random sampling. Both subsets originated from the same study population and four participating hospitals and had identical eligibility criteria, outcome definitions, and candidate predictors. Thus, the test set represented an internally held-out sample rather than an external validation dataset. This resulted in 572 training and 144 test observations. The training set included 264 (46.2%) participants without HF and 308 (53.8%) with HF, while the test set included 66 (45.8%) without HF and 78 (54.2%) with HF. The test set was reserved exclusively for final evaluation and was not used for preprocessing, feature selection, model fitting, hyperparameter tuning, or model selection..

### Data pre-processing and feature selection

To minimize information leakage, all preprocessing and feature-selection procedures were performed using training data only. ReliefF was used to rank candidate predictors according to their relevance for HF classification. The algorithm estimates predictor importance by comparing observations with neighboring instances from the same and different outcome classes, assigning higher weights to more discriminative predictors.

Feature selection was conducted separately within each training fold during cross-validation, ensuring that validation observations were not used to estimate feature relevance. The number of retained predictors (k) was treated as a hyperparameter and optimized during model development, allowing the set of selected predictors to vary across folds.

### Candidate machine-learning algorithms

Seven classification algorithms were evaluated: logistic regression (LR), radial basis function support vector machine (-SVM), k-nearest neighbors (KNN), random forest (RF), random under-sampling boosting (RUSBoost), histogram-based gradient boosting (HGB), and extremely randomized trees (Extra Trees (ET)). These algorithms represent statistical, kernel-based, distance-based, and tree-based ensemble approaches.

### Model development and evaluation

The dataset was stratified into training (n = 572; 80%) and independent test (n = 144; 20%) sets. The test set was reserved exclusively for final model evaluation and was not used for preprocessing, feature selection, hyperparameter tuning, or model selection.

The seven candidate algorithms were initially compared using stratified 10-fold cross-validation within the training set. Preprocessing and ReliefF feature selection were performed independently within each training fold to minimize information leakage. Model performance was assessed using accuracy, precision, sensitivity (recall), F1-score, area under the receiver operating characteristic curve (ROC-AUC), and area under the precision–recall curve (PR-AUC). The three algorithms with the highest mean cross-validated ROC-AUC were selected for hyperparameter optimization. Randomized search with stratified five-fold cross-validation was then conducted separately for each selected algorithm, evaluating 25 randomly sampled parameter combinations per algorithm using ROC-AUC as the optimization criterion. Algorithm-specific search spaces covered model complexity, tree and boosting parameters, learning rates, feature sampling, regularization, and class-weighting strategies, as applicable. For the radial basis function support vector machine (RBF-SVM), the search included the regularization parameter (C), kernel coefficient (gamma), and class weighting. The best configuration for each algorithm was selected based on mean cross-validated ROC-AUC.

Data from the four participating hospitals were pooled for model development and evaluation. The training and test sets were generated through participant-level stratified random sampling rather than hospital-level partitioning. Hospital-level clustering was not explicitly accounted for, and between-hospital heterogeneity in model performance was not assessed; therefore, no hospital-specific performance estimates were generated.

### Final model selection and fitting

The final model was selected based on the highest mean cross-validated ROC-AUC among the three tuned candidate algorithms. The best-performing model configuration was selected and retained for final evaluation. Its hyperparameters included the number of trees, maximum tree depth, minimum samples required for node splitting, minimum samples per leaf, and the feature-sampling strategy, as applicable. No class weighting or bootstrap sampling was applied. ReliefF retained the 20 highest-ranked predictors using 10 neighbors. The selected model and its feature-selection procedure were subsequently refitted using the complete training dataset before evaluation on the independent test set. No model updating, recalibration, or further refitting was performed after test-set evaluation.

### Independent test-set evaluation

Following model development and selection, all seven model specifications were refitted using the complete training dataset (n = 572) and evaluated once on the independent test set (n = 144). The three tuned models were fitted using their respective optimized configurations, while the remaining four used their prespecified configurations. The test set was not used for preprocessing, feature selection, hyperparameter tuning, or model selection.

For each model, predicted probabilities of documented HF status were generated and converted into binary classifications using a conventional, prespecified probability threshold of 0.50, which was not optimized using the test set. Participants with predicted probabilities ≥0.50 were classified as having documented HF, while those with probabilities <0.50 were classified as not having documented HF. Performance was assessed using accuracy, sensitivity, specificity, precision (positive predictive value), negative predictive value, F1-score, ROC-AUC, PR-AUC, and balanced accuracy. Confusion matrices were also generated to summarize true-positive, true-negative, false-positive, and false-negative classifications. The binary classifications represented predicted documented HF status and were not intended to establish a clinical diagnosis.

### Bootstrap confidence intervals

Uncertainty around the performance estimates of the final Extra Trees model was assessed using nonparametric bootstrap resampling of the independent test set. A total of 5,000 bootstrap samples were generated by resampling the 144 test observations with replacement. For each bootstrap sample, accuracy, precision, sensitivity, specificity, negative predictive value, F1-score, ROC-AUC, PR-AUC, and Brier score were calculated using the fixed predictions and predicted probabilities from the final model. Percentile-based 95% confidence intervals were obtained from the 2.5^th^ and 97.5^th^ percentiles of the bootstrap distributions. Bootstrap resampling was used only to quantify uncertainty and did not involve model refitting, feature selection, hyperparameter tuning, or model selection.

### Calibration assessment

Calibration of the final Extra Trees model was assessed using its predicted probabilities on the independent test set. Calibration was evaluated using the Brier score, calibration intercept, and calibration slope. The calibration intercept was estimated from a logistic regression model of the observed outcome on the logit-transformed predicted probabilities, with an ideal value of 0. The calibration slope was obtained from the corresponding regression coefficient, with an ideal value of 1. A calibration table was generated by grouping predicted probabilities into 10 intervals and comparing the mean predicted probability with the observed proportion of HF in each interval. A calibration plot was used to display agreement between predicted and observed probabilities, with the 45° line representing perfect calibration

### Model interpretability and feature-importance

The final Extra Trees model was interpreted using permutation importance and Shapley additive explanations (SHAP). Permutation importance was assessed on the independent test set using ROC-AUC as the performance measure. Each predictor was randomly permuted while the other predictors were left unchanged, and the resulting decrease in ROC-AUC was used as the measure of importance. Each predictor was permuted 30 times using a fixed random seed. Predictors were ranked according to their mean decrease in ROC-AUC, with larger decreases indicating greater contribution to model discrimination. The 20 highest-ranked predictors were displayed graphically.

The SHAP was additionally used to examine the contribution of individual predictors to model predictions. SHAP values were calculated for the final model and summarized using a beeswarm plot. Predictors were ranked according to their mean absolute SHAP values, with larger values indicating greater overall influence on model predictions. The 15 most influential predictors were displayed in the beeswarm plot, showing the magnitude and direction of their contributions to individual predictions. Concise labels were used for graphical presentation while the original variable definitions were retained for reporting.

### Decision curve

Decision-curve analysis was performed on the independent test set for all seven evaluated models to assess their potential clinical utility across a range of threshold probabilities. Net benefit was calculated at threshold probabilities from 0.01 to 0.98 and compared with the strategies of treating all individuals and treating none. The analysis used the fixed predicted probabilities generated by each model on the independent test set; no recalibration or threshold optimization was performed.

### Software and computational environment

All analyses were conducted in Python 3.12.7 within the Anaconda distribution version 2.6.6 using a Jupyter Notebook environment. Scikit-learn was used for ML model development, cross-validation, hyperparameter tuning, and performance evaluation. Scikit-rebate was used for ReliefF-based feature selection, and SHAP was used for model interpretation. Decision-curve analysis was implemented using a custom Python function to calculate net benefit across the specified threshold probabilities

### Ethical clearance

The study was approved by the Institutional Review Board of Hawassa University, College of Medicine and Health Sciences (Reference No. IRB/405/16; approval date: 23 December 2024). Permission was also obtained from the participating hospitals. Written informed consent was obtained from all study participants before data collection. The study was conducted in accordance with relevant ethical principles and guidelines.

### Reporting guideline

This study was reported in accordance with the Transparent Reporting of a Multivariable Prediction Model for Individual Prognosis or Diagnosis (TRIPOD+AI) 2024 guideline for clinical prediction models developed using machine-learning methods [25]. The completed TRIPOD+AI checklist is provided in Supplementary File 1.

## Results

### Dataset characteristics and outcome distribution

The ML analysis included 716 observations and 40 candidate predictor features, with HF status (heartFDx) specified as the binary target variable (0 = no HF; 1 = HF). Overall, 386 (53.9%) were classified as having HF (Table 1).

**Table 1.** The dataset characteristics and outcome distribution of the current study.

| **Characteristic** | **n (%)** |
|---|---|
| Total observations | 716 |
| Candidate predictor features | 40 |
| Prediction task | Binary classification |
| No HF (class 0) | 330 (46.1) |
| HF (class 1) | 386 (53.9) |
| Positive-class prevalence | 53.9% |
| Training set | 572 (80.0) |
| *No HF* | 264 (46.2) |
| *HF* | 308 (53.8) |
| Independent test set | 144 (20.0) |
| *No HF* | 66 (45.8) |
| *HF* | 78 (54.2) |

The dataset was divided using a stratified 80:20 split into a training set of 572 observations (80.0%) and an independent held-out test set of 144 observations (20.0%). The training set comprised 264 observations without HF (46.2%) and 308 with HF (53.8%), whereas the test set comprised 66 (45.8%) and 78 (54.2%), respectively. Thus, the outcome distribution was similar across the training and test sets (Table 1).

### Initial comparison of candidate models

Seven ML classifiers were compared using stratified 10-fold cross-validation within the training dataset only: LR, RBF-SVM, RF, RUSBoost, HGB, ET, and KNN. Performance was assessed using accuracy, precision, recall, F1-score, ROC-AUC, and PR-AUC (Table 2).

**Table 2.** Initial stratified 10-fold cross-validated performance of the candidate ML models.

| Model | Accuracy | Precision | Recall | F1-score | ROC-AUC | PR-AUC |
|---|---|---|---|---|---|---|
| **Extra trees** | **0.892** | **0.921** | **0.873** | **0.897** | **0.953** | **0.966** |
| Random forest | 0.883 | 0.914 | 0.864 | 0.888 | 0.948 | 0.958 |
| SVM | 0.883 | 0.932 | 0.844 | 0.886 | 0.943 | 0.961 |
| Logistic regression | 0.867 | 0.900 | 0.847 | 0.873 | 0.940 | 0.957 |
| RUSBoost | 0.872 | 0.927 | 0.828 | 0.875 | 0.936 | 0.952 |
| Gradient Boosting | 0.864 | 0.878 | 0.867 | 0.873 | 0.936 | 0.953 |
| KNN | 0.822 | 0.848 | 0.815 | 0.831 | 0.918 | 0.934 |

Extra trees showed the highest initial cross-validated ROC-AUC (0.953) and PR-AUC (0.966), with an accuracy of 0.892, precision of 0.921, recall of 0.873, and F1-score of 0.897. Random forest ranked second by ROC-AUC (0.948), followed by SVM (0.943). Logistic regression, RUSBoost, HGB, and KNN had progressively lower ROC-AUC values of 0.940, 0.936, 0.936, and 0.918, respectively.

The corresponding receiver ROC and precision–recall curves from the initial cross-validation are shown in Fig 1.

**Fig 1.**
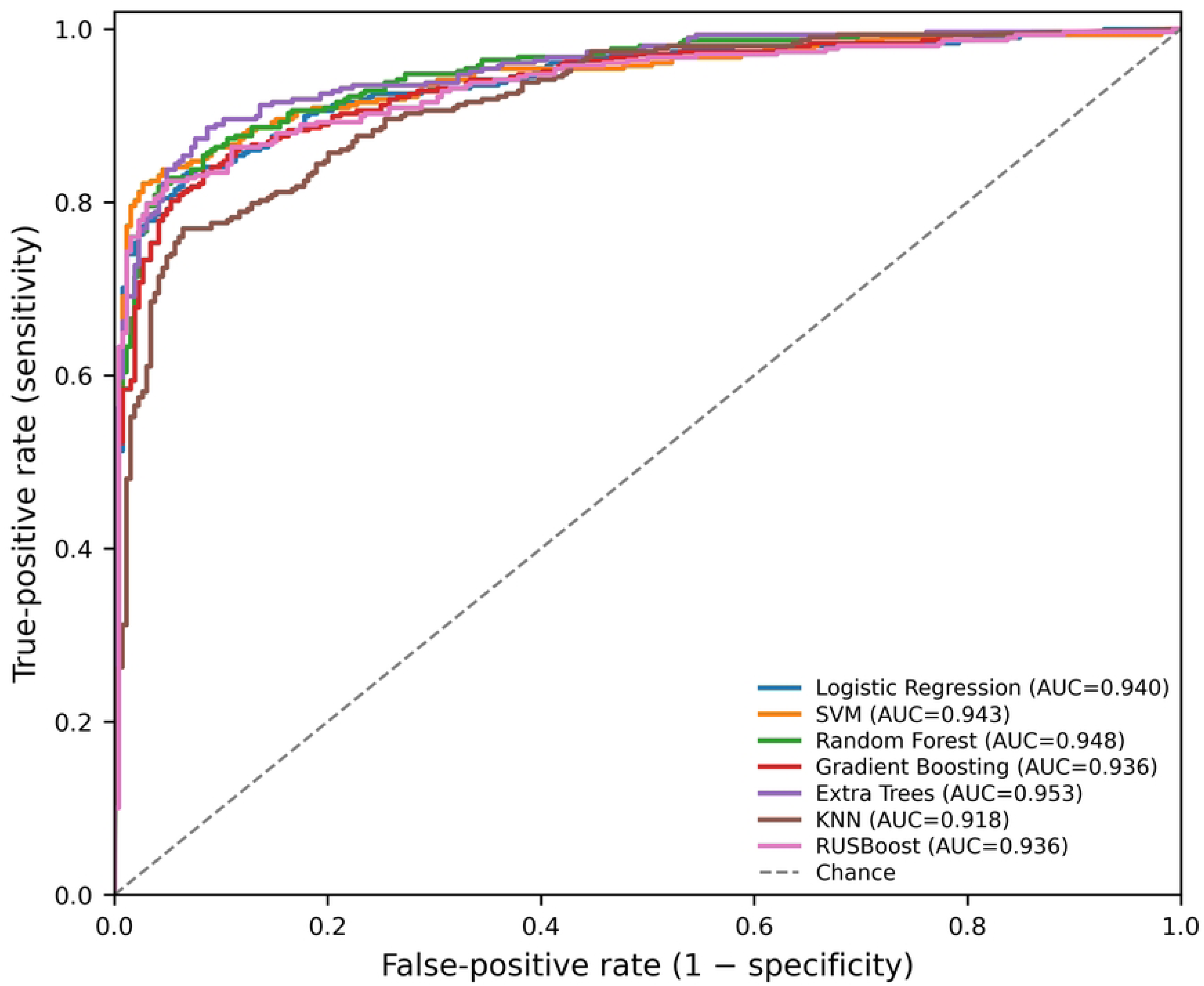
Initial cross-validated discrimination of the seven candidate models generated from out-of-fold predicted probabilities during stratified 10-fold cross-validation in the training dataset (n = 572)

### Hyperparameter optimization and model selection

The three leading models based on initial cross-validated performance—ET, RF, and SVM— were retained for hyperparameter optimization. Randomized searches evaluated 25 randomly sampled hyperparameter combinations for each model using stratified five-fold cross-validation within the training dataset. The ROC-AUC was used as the optimization criterion, and the independent test set was not used during hyperparameter tuning or model selection.

After optimization, ET achieved the highest mean five-fold cross-validated ROC-AUC (0.955). The optimal ET configuration comprised 300 trees, a maximum tree depth of 10, a minimum of five observations required for node splitting, a minimum of two observations per terminal leaf, square-root feature selection at each split, and no class weighting. The ReliefF feature-selection procedure retained 20 predictors using 10 neighbors (Table 3).

**Table 3.**
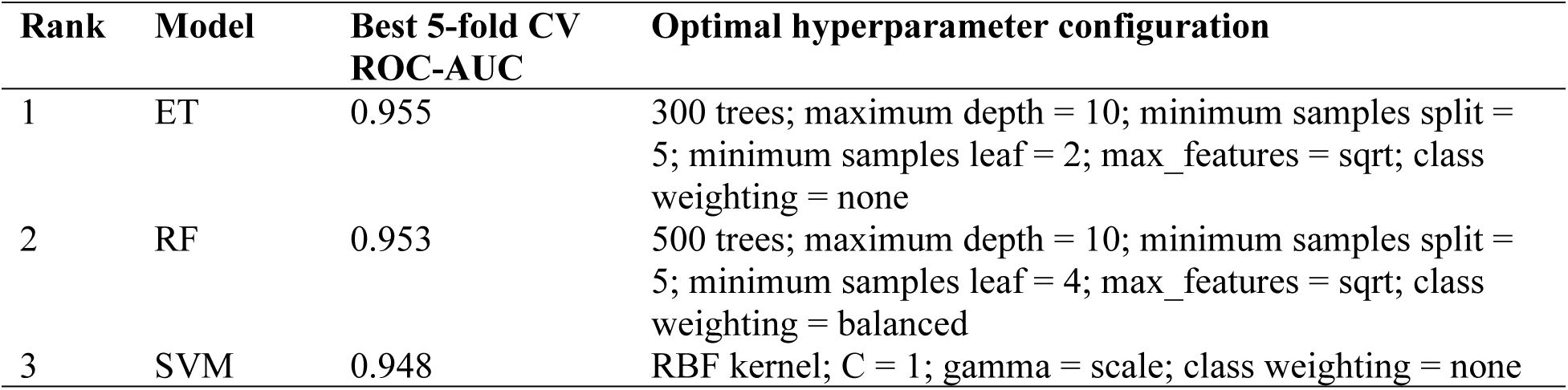
Hyperparameter-tuned cross-validated performance and optimal hyperparameter configurations of the three leading models.

Based on the highest tuned cross-validated ROC-AUC, ET was selected as the final model. The final pipeline comprised ReliefF feature selection retaining 20 predictors followed by an ET classifier using the selected hyperparameters. This pipeline was refitted using all 572 observations in the training dataset before evaluation on the independent test set.

### Independent test-set performance of the candidate models

All seven model specifications were subsequently refitted using the complete training dataset (n=572) and evaluated once on the independent held-out test set (n=144). Extra Trees achieved the highest ROC-AUC (0.954) and PR-AUC (0.966), followed by RUSBoost and RF, each with a ROC-AUC of 0.949 (Table 4).

**Table 4.** Performance of the seven model specifications on the independent test set.

| **Model** | **Accuracy** | **Sensitivity** | **Specificity** | **F1** | **ROC-AUC** | **PR-AUC** |
|---|---|---|---|---|---|---|
| ET | 0.903 | 0.872 | 0.939 | 0.907 | 0.954 | 0.966 |
| RUSBoost | 0.889 | 0.821 | 0.970 | 0.889 | 0.949 | 0.964 |
| RF | 0.889 | 0.846 | 0.939 | 0.892 | 0.949 | 0.964 |
| HGB | 0.861 | 0.859 | 0.864 | 0.870 | 0.946 | 0.962 |
| SVM | 0.882 | 0.872 | 0.894 | 0.889 | 0.945 | 0.957 |
| LR | 0.910 | 0.859 | 0.970 | 0.912 | 0.938 | 0.959 |
| KNN | 0.840 | 0.821 | 0.864 | 0.848 | 0.930 | 0.942 |

### Final ET model performance

The final ET model correctly classified 130 of 144 observations (62 true negatives, 68 true positives, four false positives, and 10 false negatives), achieving an accuracy of 0.903 (95% CI: 0.854–0.944), ROC-AUC of 0.954 (95% CI: 0.918–0.982), PR-AUC of 0.966 (95% CI: 0.938– 0.987), and Brier score of 0.082 (95% CI: 0.057–0.110) (Table 5). Confidence intervals were estimated using 5,000 non-parametric bootstrap replicates (Table 5).

**Table 5.** Performance of the final ET model on the independent test set.

| **Metric** | **Estimate** | **95% CI** |
|---|---|---|
| Accuracy | 0.903 | 0.854–0.944 |
| Precision | 0.944 | 0.887–0.987 |
| Sensitivity | 0.872 | 0.793–0.941 |
| Specificity | 0.939 | 0.875–0.986 |
| Negative predictive value (NPV) | 0.861 | 0.773–0.935 |
| F1-score | 0.907 | 0.853–0.951 |
| ROC-AUC | 0.954 | 0.918–0.982 |
| PR-AUC | 0.966 | 0.938–0.987 |
| Brier score | 0.082 | 0.057–0.110 |
| True negatives | 62 | — |
| False positives | 4 | — |
| False negatives | 10 | — |
| True positives | 68 | — |

### Discrimination

The final ET model showed strong discrimination on the independent test set, with a ROC-AUC of 0.954 (95% CI: 0.918–0.982) and PR-AUC of 0.966 (95% CI: 0.938–0.987) (Table 5, Fig 2).

**Fig 2.**
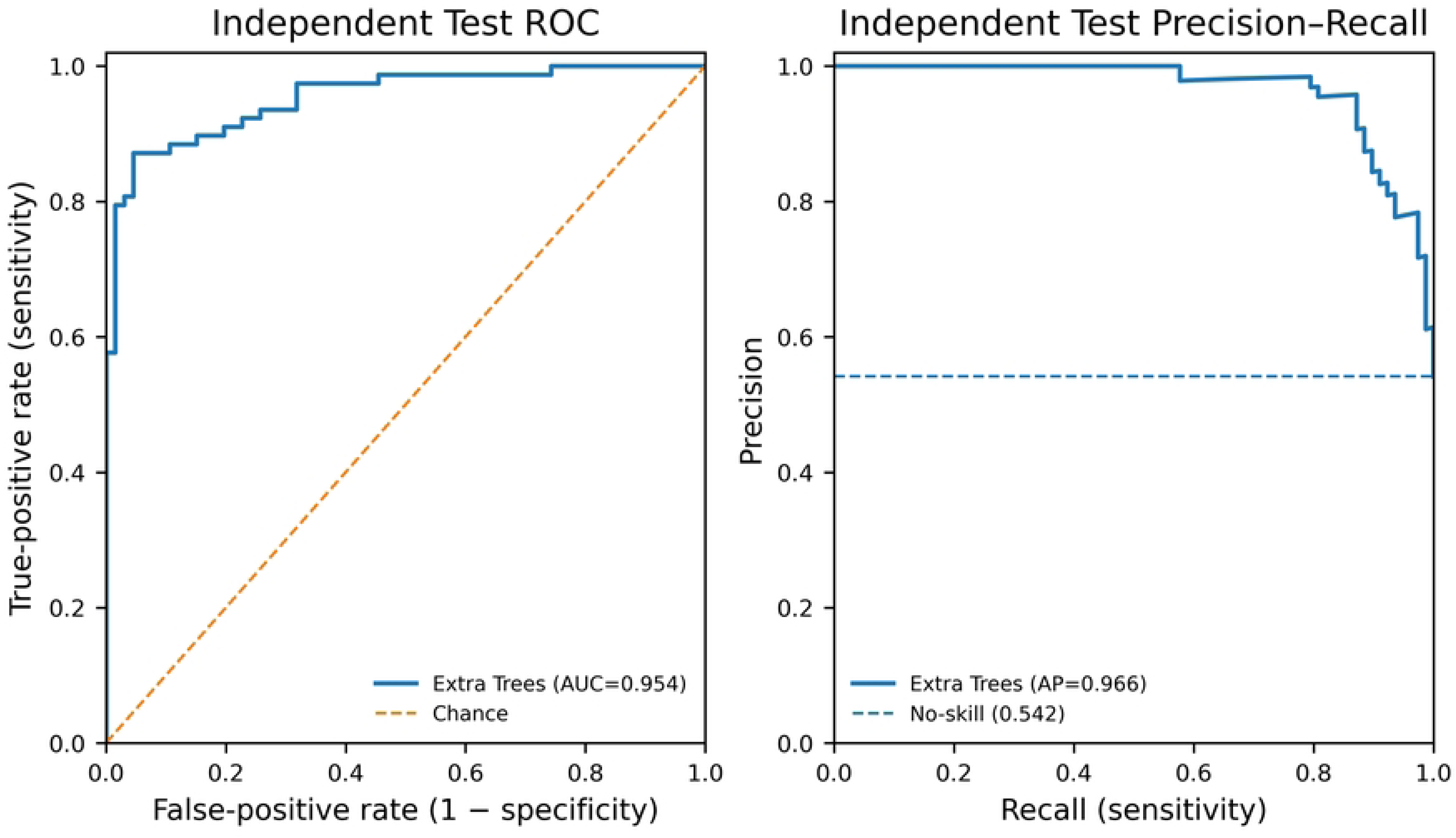
ROC and PR curves of the final ET model which evaluated on the independent held-out test set (n=144).

### Classification matrix

At the classification threshold used for model evaluation, the final ET model correctly classified 62 of 66 observations without HF and 68 of 78 observations with HF. Four observations without HF were classified as having HF, while 10 observations with HF were classified as not having HF (Fig 3).

**Fig 3.**
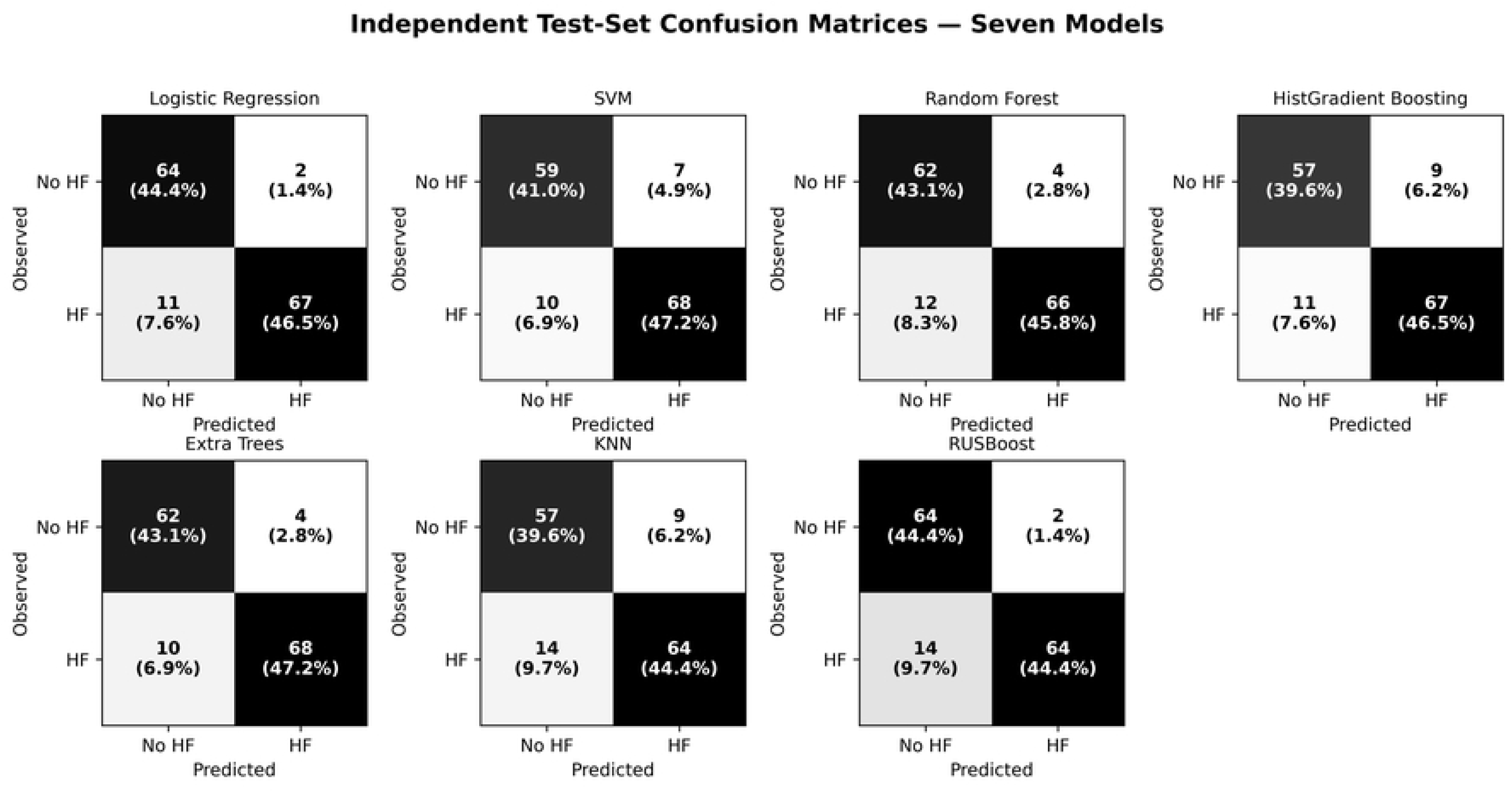
Confusion matrix of the seven models on the independent test set (n=144).

### Decision-curve

Decision-curve analysis demonstrated variation in net benefit across threshold probabilities for the seven evaluated models. The ET model provided positive net benefit across the reported range of threshold probabilities, relative to the strategies of treating all participants or treating none (Fig 4).

**Fig 4.**
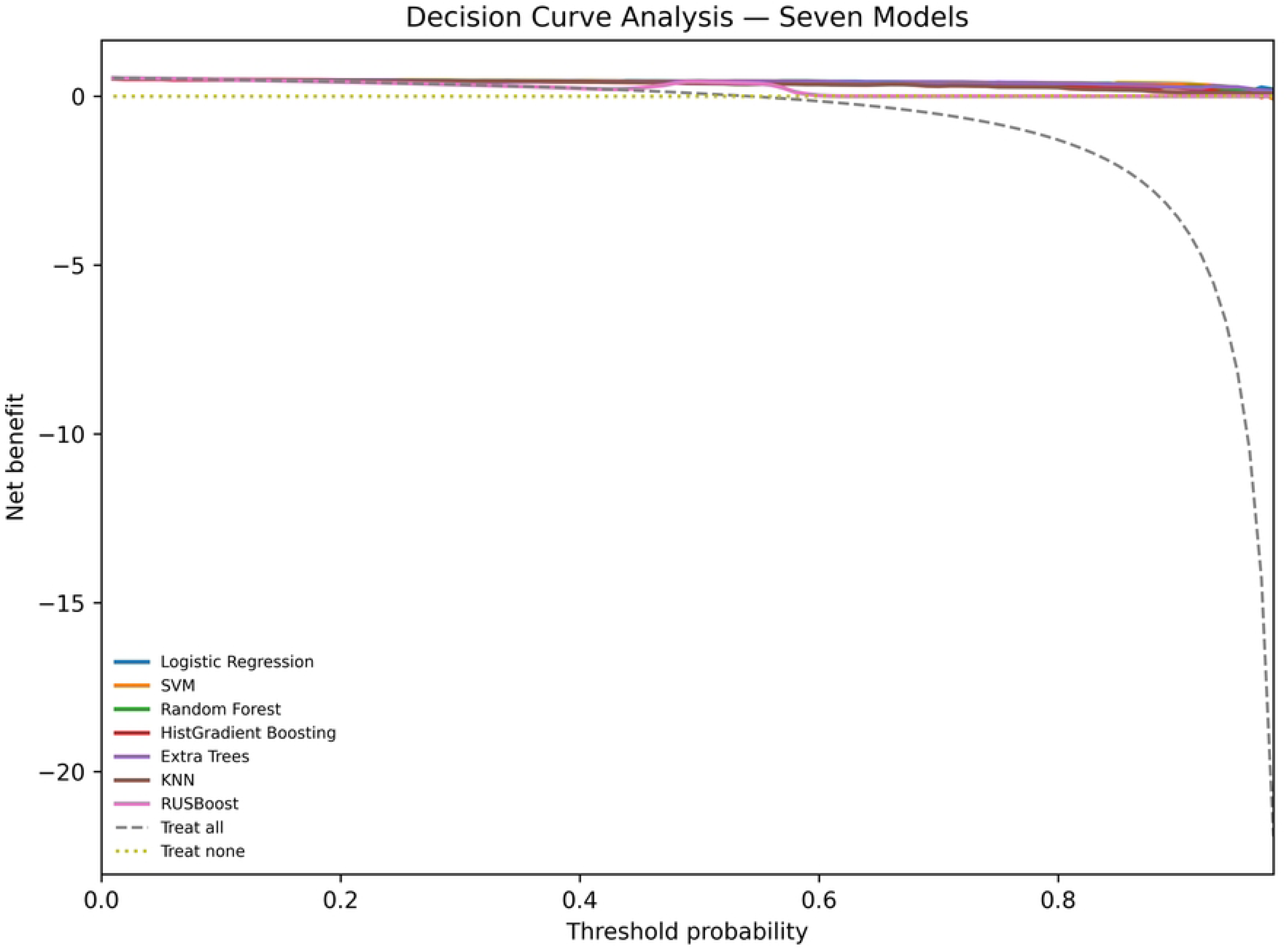
Decision-curve analysis of the seven model specifications on the independent test set.

### Calibration

The final ET model had a Brier score of 0.082 (95% CI: 0.057–0.110), indicating the overall accuracy of its predicted probabilities. The calibration intercept was 0.025, while the calibration slope was 1.352. Predicted and observed risks were closely aligned at the lowest and highest predicted-risk ranges: mean predicted and observed probabilities were 0.046 and 0.045, respectively, at the lowest range and 0.966 and 0.980, respectively, at the highest range. Greater discrepancies were observed in some intermediate probability ranges, although interpretation of these groups was limited by their smaller numbers of observations (Table 6)

**Table 6.** Calibration performance of the seven model specifications on the independent test set.

| Model | Brier score | Calibration intercept | Calibration slope |
|---|---|---|---|
| LR | 0.085 | −0.033 | 1.073 |
| SVM | 0.084 | −0.102 | 0.904 |
| RF | 0.089 | 0.111 | 1.486 |
| HGB | 0.106 | −0.066 | 0.342 |
| **ET** | **0.082** | **0.025** | **1.352** |
| KNN | 0.112 | 0.135 | 0.963 |
| RUSBoost | 0.210 | 0.175 | 15.694 |

### Predictor importance

The final ET model incorporated 20 predictors selected using ReliefF. Permutation importance was quantified as the mean decrease in ROC-AUC after randomly permuting each predictor.

Prior history of heart attack or stroke was the most influential predictor, with a mean decrease in ROC-AUC of 0.2266 (SD 0.0328). This was substantially larger than the corresponding decreases associated with sleep disorder (0.0189, SD 0.0050), current medication use for blood pressure or cholesterol (0.0102, SD 0.0064), systolic blood pressure (0.0080, SD 0.0032), and age (0.0076, SD 0.0026). The remaining predictors had relatively small decreases in ROC-AUC, and alcohol consumption and family history of cardiovascular disease produced no decrease in ROC-AUC in the permutation analysis (Table 7).

**Table 7.** Permutation importance of predictors retained in the final ET model.

| **Rank** | **Predictor** | **Mean decrease in ROC-AUC** | **SD** |
|---|---|---|---|
| 1 | Prior heart attack/stroke | 0.2266 | 0.0328 |
| 2 | Sleep disorder | 0.0189 | 0.0050 |
| 3 | Blood pressure/cholesterol medication | 0.0102 | 0.0064 |
| 4 | Systolic blood pressure | 0.0080 | 0.0032 |
| 5 | Age | 0.0076 | 0.0026 |
| 6 | Sex | 0.0059 | 0.0032 |
| 7 | Self-rated health | 0.0055 | 0.0055 |
| 8 | Benzathine penicillin history | 0.0055 | 0.0023 |
| 9 | Prior CVD diagnosis | 0.0051 | 0.0031 |
| 10 | Educational status | 0.0051 | 0.0021 |
| 11 | Marital status | 0.0050 | 0.0029 |
| 12 | Region | 0.0041 | 0.0026 |
| 13 | Moderate physical activity | 0.0027 | 0.0021 |
| 14 | Current stress/depression | 0.0023 | 0.0025 |
| 15 | Grain consumption | 0.0023 | 0.0014 |
| 16 | Employment status | 0.0015 | 0.0022 |
| 17 | Previous rheumatic fever | 0.0006 | 0.0011 |
| 18 | Religion | 0.0002 | 0.0022 |
| 19 | Alcohol consumption | 0.0000 | 0.0000 |
| 20 | Family CVD history | 0.0000 | 0.0000 |

### SHAP-based model interpretation

Global SHAP analysis identified prior history of heart attack or stroke as the most influential predictor, with a mean absolute SHAP value of 0.2308. The next most influential predictors were current use of medication for blood pressure or cholesterol (0.0845), sleep disorders (0.0493), medication adherence (0.0404), self-rated health status (0.0289), previous cardiovascular disease diagnosis (0.0259), and region (0.0181). The remaining predictors had smaller mean absolute SHAP values (Table 8, Fig 5).

**Fig 5.**
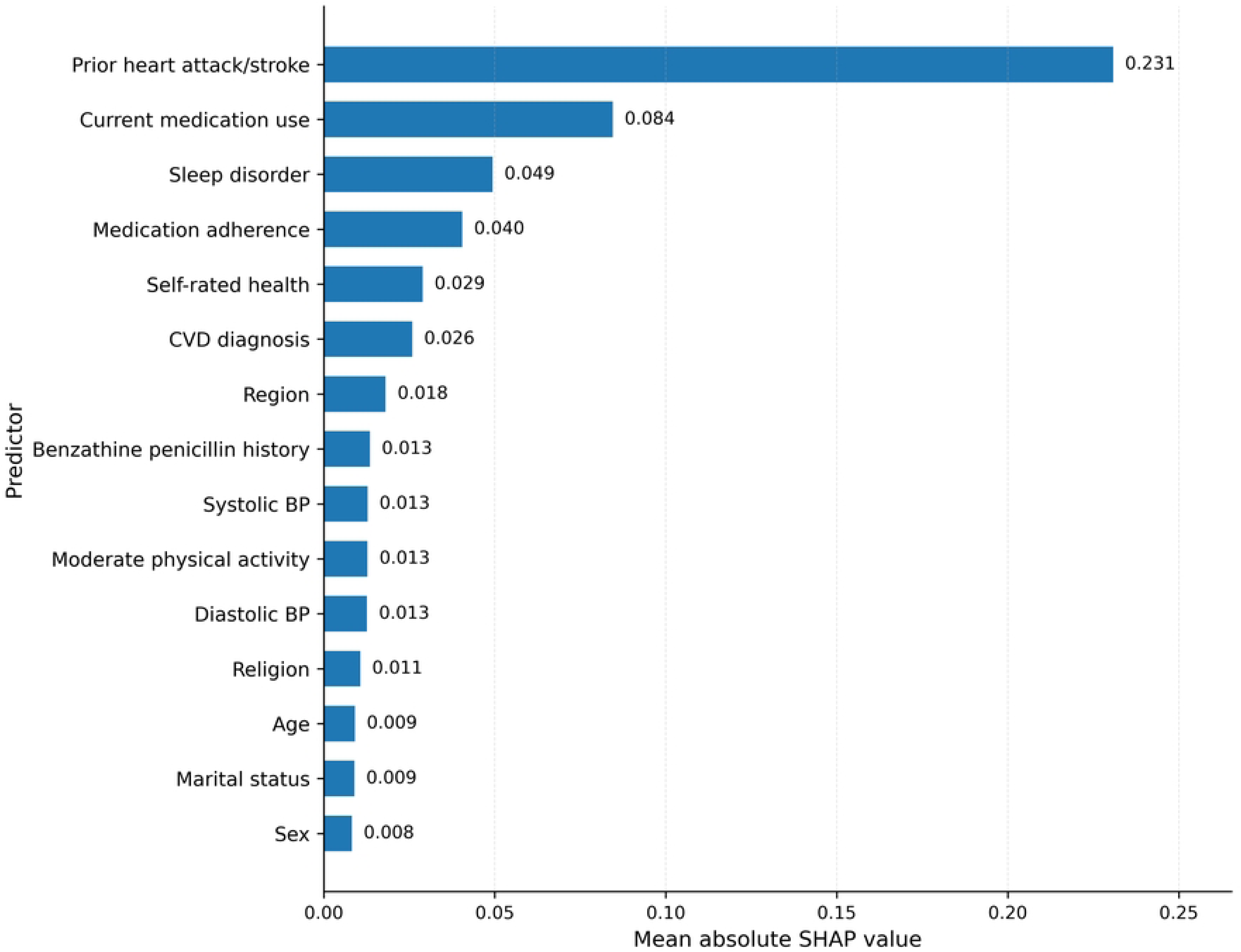
Global SHAP-based feature importance of the final ET model.

**Table 8.**
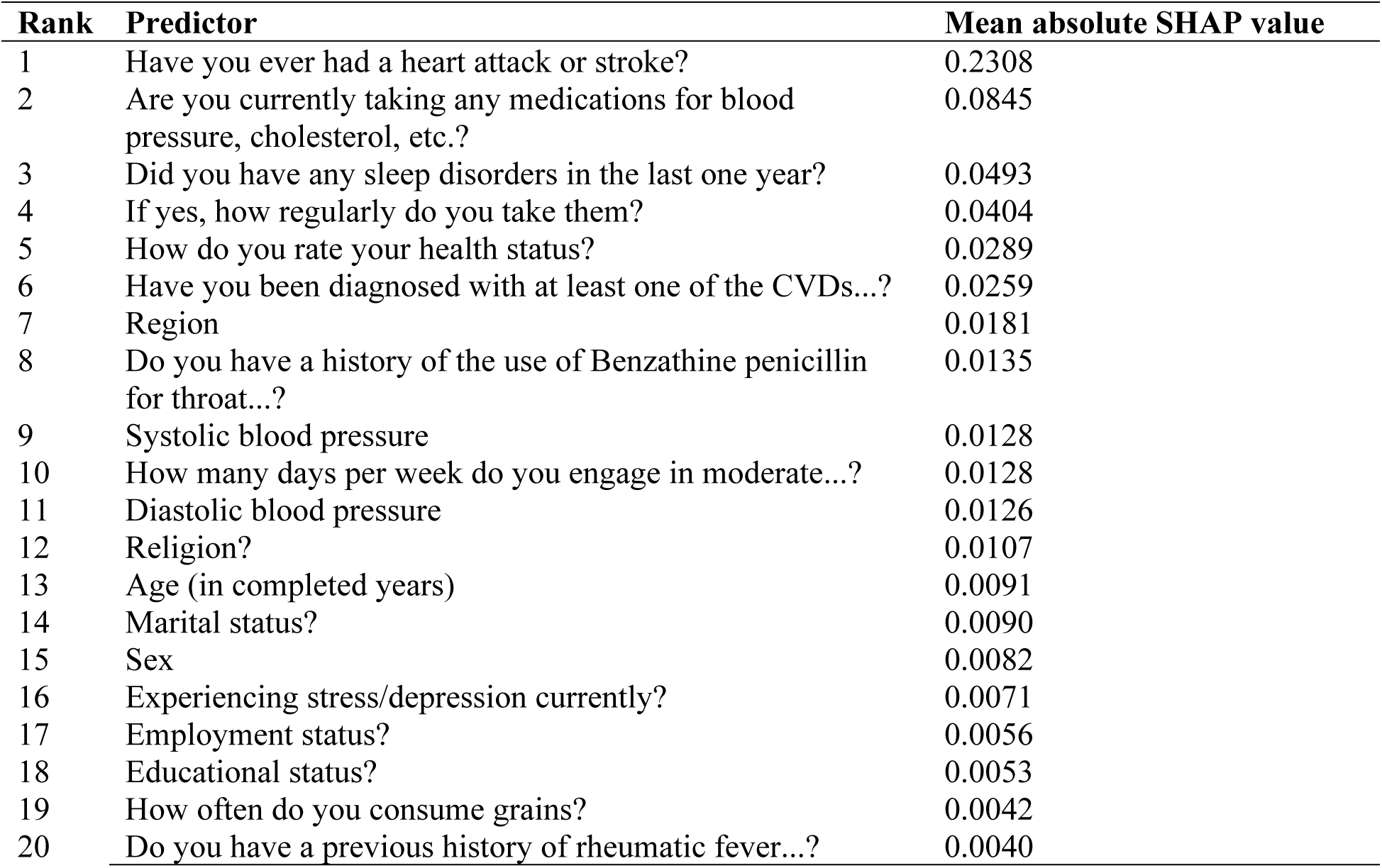
Global SHAP importance of predictors retained in the final Extra Trees model.

| **Rank** | **Predictor** | **Mean absolute SHAP value** |
|---|---|---|
| 1 | Have you ever had a heart attack or stroke? | 0.2308 |
| 2 | Are you currently taking any medications for blood pressure, cholesterol, etc.? | 0.0845 |
| 3 | Did you have any sleep disorders in the last one year? | 0.0493 |
| 4 | If yes, how regularly do you take them? | 0.0404 |
| 5 | How do you rate your health status? | 0.0289 |
| 6 | Have you been diagnosed with at least one of the CVDs...? | 0.0259 |
| 7 | Region | 0.0181 |
| 8 | Do you have a history of the use of Benzathine penicillin for throat...? | 0.0135 |
| 9 | Systolic blood pressure | 0.0128 |
| 10 | How many days per week do you engage in moderate...? | 0.0128 |
| 11 | Diastolic blood pressure | 0.0126 |
| 12 | Religion? | 0.0107 |
| 13 | Age (in completed years) | 0.0091 |
| 14 | Marital status? | 0.0090 |
| 15 | Sex | 0.0082 |
| 16 | Experiencing stress/depression currently? | 0.0071 |
| 17 | Employment status? | 0.0056 |
| 18 | Educational status? | 0.0053 |
| 19 | How often do you consume grains? | 0.0042 |
| 20 | Do you have a previous history of rheumatic fever...? | 0.0040 |

The SHAP beeswarm plot further illustrates the magnitude and direction of individual predictor contributions across observations. Predictors farther from zero contributed more strongly to individual predictions, while the distribution of SHAP values illustrates heterogeneity in predictor contributions across observations (Fig 6).

**Fig 6.**
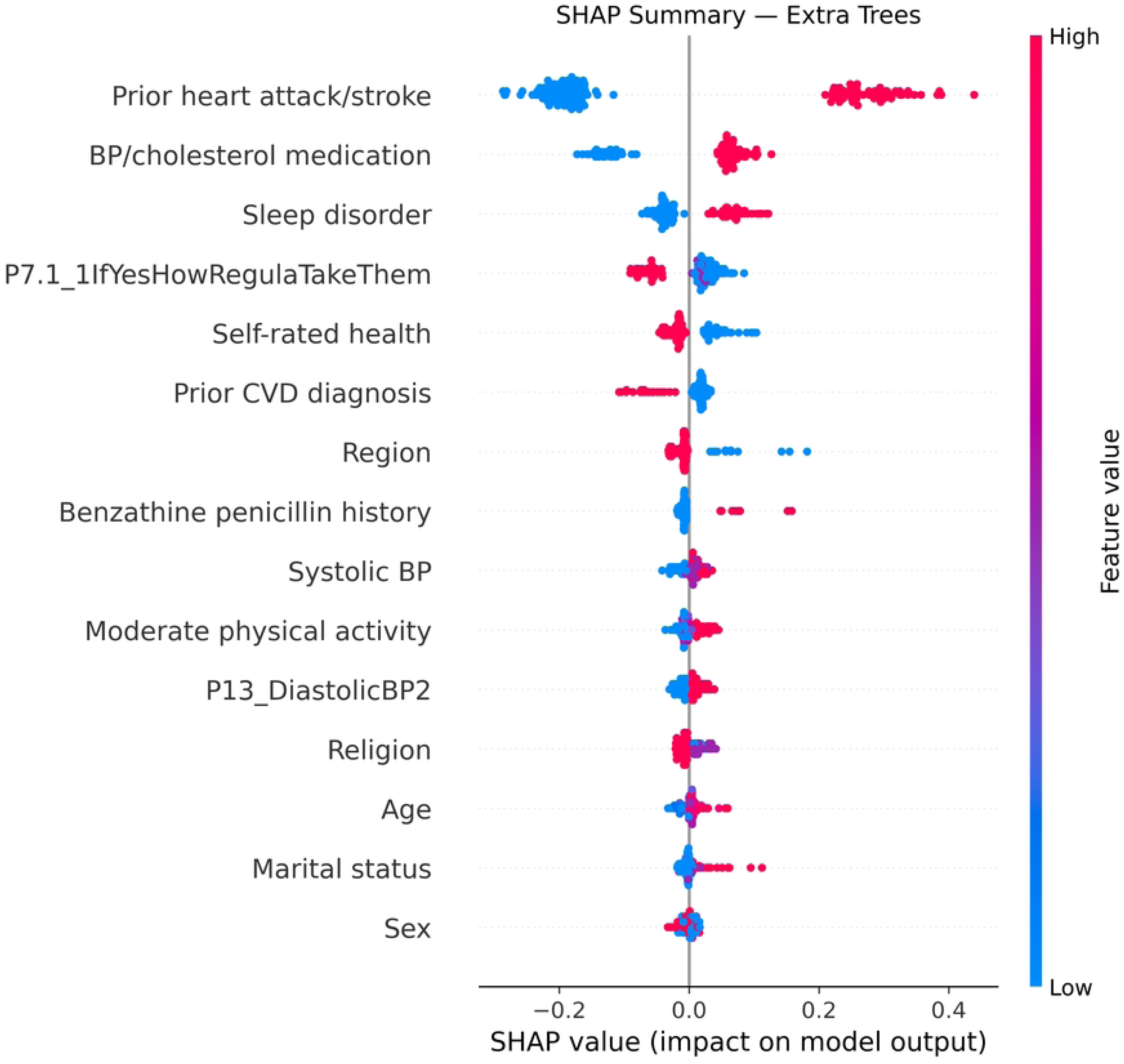
SHAP beeswarm plot of the final ET model in which 15 most influential input features are ranked mean absolute SHAP value.

SHAP dependence plots were used to further examine the contribution of the three most influential predictors: prior heart attack or stroke, sleep disorder, and current blood pressure/ cholesterol medication use. Prior heart attack or stroke showed the largest range of SHAP contributions, whereas sleep disorder and medication use showed smaller but heterogeneous contributions across observations. The corresponding dependence plots are presented in Fig 7.

**Fig 7.**
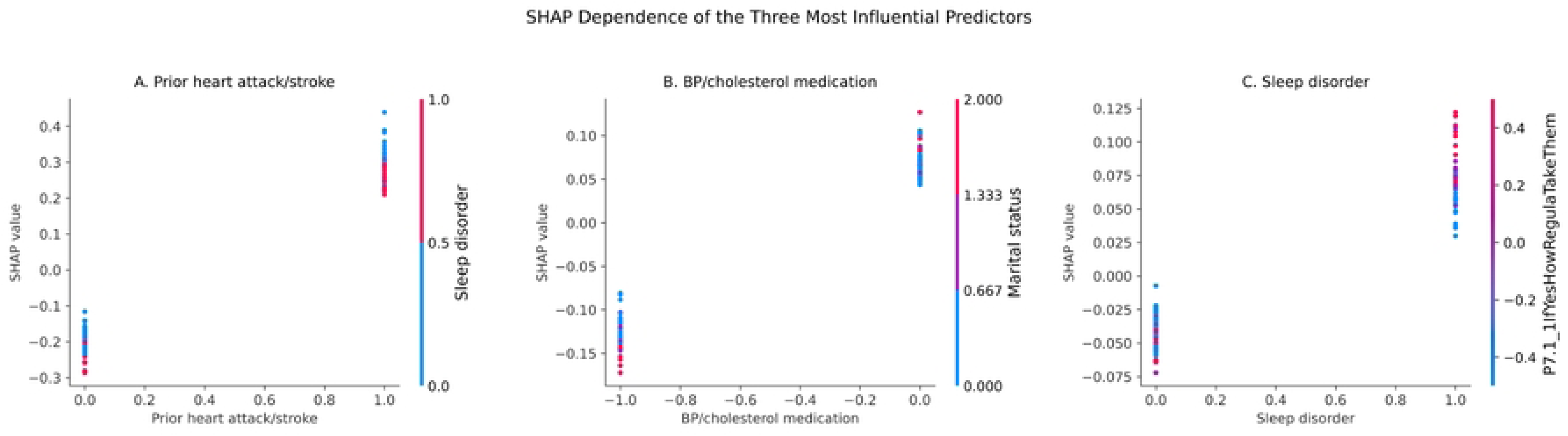
SHAP dependence plots for the three most influential predictors. Panels show the relationship between predictor values and SHAP values in the final Extra Trees model: (A) prior heart attack or stroke, (B) sleep disorder, and (C) blood pressure/cholesterol medication use. SHAP values above zero indicate contributions toward the positive HF prediction and values below zero indicate contributions toward the negative prediction.

## Discussion

In this multicenter, cross-sectional ML study of 716 patients attending four public hospitals in Sidama region, Ethiopia, the ET classifier showed the highest discrimination for distinguishing participants with a documented diagnosis of HF from those without. On the independent held-out test set, the model achieved a ROC-AUC of 0.954 and a PR-AUC of 0.966, with 90.3% accuracy, 87.2% sensitivity and 93.9% specificity. Its Brier score was 0.082. Prior history of heart attack or stroke was the most influential predictor in both permutation-importance and SHAP analyses. These findings indicate that combinations of demographic and clinical characteristics can distinguish patients with and without documented HF in this hospital-based sample. However, the model’s performance reflects internal classification of the two clinically defined groups and does not establish its ability to diagnose previously unrecognized HF or replace standard diagnostic investigations.

Extra trees showed high discrimination across cross-validation, hyperparameter optimization and held-out testing (ROC-AUC 0.953, 0.955 and 0.954, respectively). However, its advantage over other algorithms was modest. Random forest and RUSBoost achieved test ROC-AUCs of 0.949, while LR had slightly higher accuracy (91.0%) and F1-score (0.912) than ET (90.3% and 0.907, respectively). Therefore, no single algorithm was superior across all performance measures. The preferred model depends on the intended application and the balance between discrimination, classification performance, interpretability and calibration. Moreover, because only the three leading algorithms underwent hyperparameter tuning, these comparisons should not be interpreted as a definitive ranking of fully optimized models.

The observed performance in the current study is consistent with previous applications of ML and AI for HF identification and classification using EHR data, clinical characteristics and digital diagnostic technologies [17,19–21]. However, these studies address heterogeneous tasks, including HF detection, identification of coded HF cases, prediction of readmission among patients with established HF, and detection of HF with reduced ejection fraction. Unlike these approaches, the present study integrates questionnaire-derived, demographic and clinical characteristics to distinguish patients with established HF from non-HF OPD attendees. It therefore extends the evidence on ML-based HF classification to a relatively resource-constrained African hospital context, although direct comparisons of predictive performance should be interpreted cautiously as differences in prediction targets, populations, data sources and validation designs.

The prominence of previous heart attack or stroke in both permutation and SHAP analyses is clinically plausible, as these conditions may reflect underlying cardiovascular damage and disease burden. Its higher permutation importance suggests that the model relied heavily on this information when distinguishing HF from non-HF patients. Other contributing features included antihypertensive or cholesterol-lowering medication use, sleep disorders, medication adherence, self-rated health, previous CVD diagnosis and blood pressure measurements. Together, these features may reflect cardiovascular risk, comorbidities and previous healthcare experiences.

However, as the HF participants were recruited from follow-up clinics, medication use and adherence may reflect prior diagnosis, prescribing and ongoing clinical care rather than independent early indicators of HF. Similarly, a history of heart attack or stroke may partly distinguish patients with previously recognized CVD. Although these features may be useful for identifying patients with established clinical histories who warrant further assessment, their importance should not be interpreted as evidence of causality or independent clinical effects, and it limits claims that the model supports early case-finding based on pre-diagnostic features.

The distinction between the study’s classification target and clinical diagnosis is important for interpreting its clinical implications. Heart failure diagnosis requires clinical assessment supported by objective evidence of cardiac structural or functional abnormalities. However, the current model used documented clinical diagnoses as its outcome without independently confirming HF status using a uniform reference standard. Additionally, controls were defined by the absence of a documented HF diagnosis, potentially including individuals with undiagnosed HF. This asymmetry in outcome ascertainment may have introduced misclassification, meaning that the model distinguishes documented HF cases from selected controls rather than independently diagnosing HF. Its potential role should therefore be considered as an aid to triage or prioritization for confirmatory assessment, not as a stand-alone diagnostic test. This interpretation aligns with TRIPOD+AI and PROBAST+AI recommendations on evaluating prediction models in populations and settings relevant to their intended use [25,26].

The calibration findings were less favorable than the discrimination results alone suggest. The ET model had a Brier score of 0.082 and a calibration intercept of 0.025, but its calibration slope was 1.352, indicating that predicted probabilities were insufficiently dispersed relative to observed outcomes. Thus, predicted probabilities should not yet be interpreted as well-calibrated individual risks. Furthermore, the selected sampling of HF follow-up patients and OPD controls yielded an HF proportion of 53.9%, not a population prevalence estimate. Predictive values and absolute probabilities may therefore differ across clinical and community settings, indicating the need for recalibration and validation in representative target populations before clinical application [27,28].

Decision-curve analysis suggested positive net benefit for ET across the reported threshold range, providing an assessment beyond discrimination and accuracy. However, net benefit depends on a clearly defined clinical action and thresholds reflecting the consequences of false-positive and false-negative decisions. Referral for further HF assessment may be a plausible action, but the relevant thresholds require clinical justification. Given the small held-out sample and absence of prospective evaluation, these findings should be interpreted as exploratory evidence of potential clinical utility, not proof of clinical benefit [29].

The study has several strengths. Its multicenter design included four public hospitals, providing diversity in clinical settings, and compared seven algorithms representing regression, kernel-based, distance-based and tree-ensemble approaches. The modelling workflow incorporated preprocessing and ReliefF feature selection within training folds, an independent held-out test set, and multiple complementary performance measures. Bootstrap confidence intervals, calibration, decision-curve analysis, permutation importance and SHAP provided additional assessment of model performance, potential clinical utility and interpretability. Reporting followed TRIPOD+AI [25]. Finally, the use of demographic and clinical characteristics potentially obtainable without advanced imaging enhances the relevance of the approach to resource-constrained settings.

Several limitations should be considered. First, the relatively small sample, particularly the independent test set (n = 144), limits the precision of performance and calibration estimates. Moreover, bootstrap intervals based on fixed predictions do not capture uncertainty from repeating the entire model-development process. Second, recruiting established HF cases from cardiac follow-up clinics and controls from general OPD may have introduced selection and spectrum bias, potentially inflating discrimination compared with less selected clinical populations. Third, HF status was based on documented diagnoses, and controls were not systematically assessed using a uniform reference standard to exclude HF. Fourth, the importance of prior cardiovascular events and treatment-related variables suggests that some predictors may reflect previous diagnosis and healthcare utilization. Fifth, evaluation was restricted to an internal hold-out sample from the same hospitals and study period, without external validation. Hospital-level clustering and between-hospital variation in model performance were also not assessed, limiting conclusions about transportability across facilities. Finally, the sample was insufficient for robust subgroup and fairness assessments, only the three leading algorithms underwent hyperparameter tuning, and no formal paired AUC comparisons were conducted.

Taken together, these findings demonstrate promising internal discrimination of documented HF status using routinely available demographic and clinical information in a selected hospital-based sample. The model warrants further evaluation as a potential triage or decision-support aid to identify patients requiring definitive HF assessment. However, it should not yet be used for diagnosis or population screening, or as a substitute for guideline-recommended evaluation.

External validation, assessment of predictor availability before diagnosis, setting-specific recalibration, and prospective evaluation of clinical consequences are necessary before implementation. These limitations should be considered when appraising the quality, risk of bias, and applicability of the prediction model [26–28].

### Conclusions and recommendations

The ET classifier demonstrated high discrimination between patients with documented HF and hospital OPD attendees without a documented HF diagnosis in Sidama, Ethiopia (ROC-AUC 0.954, sensitivity 87.2%, specificity 93.9%). Previous history of heart attack or stroke was the dominant contributor to model predictions. These findings suggest that routinely available demographic and clinical information may help distinguish documented HF status in the studied hospital population. However, the selected recruitment design, reliance on documented diagnoses, potential post-diagnostic predictors, small test set, and absence of external validation limit diagnostic and clinical interpretations. The model should therefore be considered a candidate triage aid requiring further validation, not a diagnostic tool or substitute for clinical assessment and echocardiography.

For potential implementation, the availability, completeness and quality of predictor information would need to be assessed at the point of care. Missing or unreliable predictor values should be identified and handled using prespecified procedures that are consistent with the model’s development pipeline, rather than assuming that all required information will be routinely available. If implemented as a clinical decision-support tool, the model would be intended to support healthcare professionals in prioritizing patients for further HF assessment, with clinical interpretation of its outputs and confirmatory diagnostic evaluation remaining the responsibility of qualified healthcare providers.

Future research should prioritize external validation in larger, prospective, geographically diverse cohorts of patients with suspected HF, using consistent clinical and objective reference standards. Studies should also assess performance after excluding potentially post-diagnostic predictors, evaluate subgroup discrimination and calibration, and establish clinically appropriate decision thresholds for actions such as echocardiography or specialist referral. Prospective impact studies should determine whether model-assisted triage improves timely diagnosis and referral without unacceptable false-positive referrals, missed cases, workload, or inequity. Transparent reporting of the model specification, preprocessing, feature definitions, and code should follow TRIPOD+AI recommendations.

## Data availability

The de-identified dataset supporting the findings of this study and the analytical scripts used for data preprocessing, feature selection, model development, hyperparameter tuning, and model evaluation are provided as Supporting Information files: S1 File (TRIPOD+AI checklist), S2 File (dataset), and S3 File (analytical code). These files will be made publicly available upon publication.

## Patient and public involvement

Patients and members of the public were not involved in the design, conduct, analysis, interpretation, or reporting of this study. Patients participated as study respondents and provided clinical and demographic information for model development and evaluation.

## List of acronyms/ abbreviations

AI: Artificial intelligence
AUC: Area under the curve
CI: Confidence interval
CVD: Cardiovascular diseases
EHR: Electronic health records
ET: Extremely randomized trees / extra trees
HF: Heart failure
HGB: Histogram-based gradient boosting
KNN: *k*-nearest neighbors
LMIC: Low- and middle-income countries
LR: Logistic regression
ML: Machine learning
NPV: Negative predictive value
OPD: Outpatient departments
PR-AUC: Precision–recall area under the curve
RBF-SVM: Radial basis function support vector machine
RF: Random forest
ROC-AUC: Receiver operating characteristic area under the curve
RUSBoost: Random under-sampling boosting
SD: Standard deviation
SHAP: Shapley additive explanations
SMOTE: Synthetic minority oversampling technique
SSA: Sub-Saharan Africa
SVM: Support vector machine
TRIPOD+AI: Transparent Reporting of a Multivariable Prediction Model for Individual Prognosis or Diagnosis

## Acknowledgement

We acknowledge the Sidama Regional Health Bureau and the participating health facilities for granting permission and facilitating the conduct of the study. We extend our appreciation to all study participants and data collectors for their valuable time, cooperation, and contributions.

## Notes

### Competing Interest Statement

The authors have declared no competing interest.

### Author Declarations

Institutional Review Board of the College of Medicine and Health Sciences, Hawassa University

## References

1. Bozkurt B, Coats AJS, Tsutsui H, Abdelhamid CM, Adamopoulos S, Albert N, et al. Universal definition and classification of heart failure: a report of the Heart Failure Society of America, Heart Failure Association of the European Society of Cardiology, Japanese Heart Failure Society and Writing Committee of the Universal Definition of Heart Failure: Endorsed by the Canadian Heart Failure Society, Heart Failure Association of India, Cardiac Society of Australia and New Zealand, and Chinese Heart Failure Association. Eur J Heart Fail. 2021;23: 352–380. doi:10.1002/ejhf.2115

2. Yan S, Deng P, Chai K, Wang S, Hu K, Yang J, et al. Global, regional, and national burden of heart failure and its risk factors between 1990 and 2021 and projections to 2050: an analysis of the global burden of disease study. Eur Heart J Qual Care Clin Outcomes. 2025;11: 1144–1154. doi:10.1093/ehjqcco/qcaf054

3. Mensah GA, Fuster V, Roth GA. A Heart-Healthy and Stroke-Free World: Using Data to Inform Global Action. J Am Coll Cardiol. 2023;82: 2343–2349. doi:10.1016/j.jacc.2023.11.003

4. McDonagh TA, Metra M, Adamo M, Gardner RS, Baumbach A, Böhm M, et al. 2021 ESC Guidelines for the diagnosis and treatment of acute and chronic heart failure. Eur Heart J. 2021;42: 3599–3726. doi:10.1093/eurheartj/ehab368

5. Heidenreich PA, Bozkurt B, Aguilar D, Allen LA, Byun JJ, Colvin MM, et al. 2022 AHA/ACC/HFSA Guideline for the Management of Heart Failure: A Report of the American College of Cardiology/American Heart Association Joint Committee on Clinical Practice Guidelines. Circulation. 2022;145: e895–e1032. doi:10.1161/CIR.0000000000001063

6. Mant J, Doust J, Roalfe A, Barton P, Cowie MR, Glasziou P, et al. Systematic review and individual patient data meta-analysis of diagnosis of heart failure, with modelling of implications of different diagnostic strategies in primary care. Health Technol Assess. 2009;13: 1–207, iii. doi:10.3310/hta13320

7. Damasceno A, Mayosi BM, Sani M, Ogah OS, Mondo C, Ojji D, et al. The causes, treatment, and outcome of acute heart failure in 1006 Africans from 9 countries. Arch Intern Med. 2012;172: 1386–1394. doi:10.1001/archinternmed.2012.3310

8. Sliwa K, Hahnle J, Stewart S, Allie S, Damasceno A, Smit D, et al. Aetiology, management, and outcomes of acute heart failure in 17 African countries (THESUS-HF II): a prospective, multicentre, observational cohort study. Lancet. 2026;408: 1186–1197. doi:10.1016/S0140-6736(26)01657-0

9. Yuyun MF, Sliwa K, Kengne AP, Mocumbi AO, Bukhman G. Cardiovascular Diseases in Sub-Saharan Africa Compared to High-Income Countries: An Epidemiological Perspective. Glob Heart. 2020;15: 15. doi:10.5334/gh.403

10. Bekele F, Sheleme T, Tsegaye T, Parameswari SA, Syed MA, Tafese L, et al. Prevalence and risk factors of mortality among heart failure patients in low resource setting hospitals: a multicenter prospective observational study. Front Cardiovasc Med. 2024;11: 1429513. doi:10.3389/fcvm.2024.1429513

11. Solela G, Yimer YS. Clinical characteristics, treatment patterns, and outcomes of hospitalized patients with acute heart failure in central Ethiopia: a retrospective observational study. BMC Cardiovasc Disord. 2024;24: 241. doi:10.1186/s12872-024-03905-z

12. Blecker S, Katz SD, Horwitz LI, Kuperman G, Park H, Gold A, et al. Comparison of Approaches for Heart Failure Case Identification From Electronic Health Record Data. JAMA Cardiol. 2016;1: 1014–1020. doi:10.1001/jamacardio.2016.3236

13. Rajkomar A, Dean J, Kohane I. Machine Learning in Medicine. N Engl J Med. 2019;380: 1347–1358. doi:10.1056/NEJMra1814259

14. Flok A, Kajüter Rodrigues P, Pohurskyy S, Teuteberg F. A systematic review of machine learning algorithms for mortality risk, readmission and phenotype prediction in patients with heart failure: exploring key data sources, input variables and outcomes. BMC Med Inform Decis Mak. 2026;26: 205. doi:10.1186/s12911-026-03560-8

15. Saqib M, Perswani P, Muneem A, Mumtaz H, Neha F, Ali S, et al. Machine learning in heart failure diagnosis, prediction, and prognosis: review. Ann Med Surg (Lond). 2024;86: 3615–3623. doi:10.1097/MS9.0000000000002138

16. Hoang MT, Aung N, Chen Y, Slabaugh G. A systematic review of multimodal machine learning models for heart failure classification and prognosis prediction. Front Cardiovasc Med. 2026;13: 1770869. doi:10.3389/fcvm.2026.1770869

17. Sumsion D, Davis E, Fernandes M, Wei R, Milde R, Veltink JM, et al. Identification of Patients With Congestive Heart Failure From the Electronic Health Records of Two Hospitals: Retrospective Study. JMIR Med Inform. 2025;13: e64113. doi:10.2196/64113

18. Raat W, Smeets M, Henrard S, Aertgeerts B, Penders J, Droogne W, et al. Machine learning optimization of an electronic health record audit for heart failure in primary care. ESC Heart Fail. 2022;9: 39–47. doi:10.1002/ehf2.13724

19. Oo MM, Gao C, Cole C, Hummel Y, Guignard-Duff M, Jefferson E, et al. Artificial intelligence-assisted automated heart failure detection and classification from electronic health records. ESC Heart Fail. 2024;11: 2769–2777. doi:10.1002/ehf2.14828

20. Okoh AK, Appiah LT, Wiafe YA, Amponsah MK, Fugar SS, Ozturk E, et al. Artificial Intelligence-Enabled Digital Auscultation for Detecting Heart Failure With Reduced Ejection Fraction in Sub-Saharan Africa: The DAMSUN-HF Study. Circulation. 2026;153: 361–363. doi:10.1161/CIRCULATIONAHA.125.078246

21. Rizinde T, Ngaruye I, Cahill ND. Comparing Machine Learning Classifiers for Predicting Hospital Readmission of Heart Failure Patients in Rwanda. J Pers Med. 2023;13: 1393. doi:10.3390/jpm13091393

22. Tegegnie AK, Tewolde K. Interpretable ensemble machine learning framework for cardiovascular disease prediction using EMR data and large language models in Ethiopia. PLOS ONE. 2026;21: e0342256. doi:10.1371/journal.pone.0342256

23. Ethiopian Statistical Services. Population and Housing Census of Ethiopia 2007. Addis Ababa, Ethiopia: Ethiopian Statistical Services; 2008. Available: https://ess.gov.et/download/population-and-housing-census-2007-national_statistical/

24. Ethiopian Statistical Services. Population Size by Sex, Area and Density by Region, Zone and Wereda: July 2024. Addis Ababa, Ethiopia; 2024. Available: https://ess.gov.et/download/projected_population-2024/

25. Collins GS, Moons KGM, Dhiman P, Riley RD, Beam AL, Calster BV, et al. TRIPOD+AI statement: updated guidance for reporting clinical prediction models that use regression or machine learning methods. BMJ. 2024;385: e078378. doi:10.1136/bmj-2023-078378

26. Moons KGM, Damen JAA, Kaul T, Hooft L, Andaur Navarro C, Dhiman P, et al. PROBAST+AI: an updated quality, risk of bias, and applicability assessment tool for prediction models using regression or artificial intelligence methods. BMJ. 2025;388: e082505. doi:10.1136/bmj-2024-082505

27. Riley RD, Archer L, Snell KIE, Ensor J, Dhiman P, Martin GP, et al. Evaluation of clinical prediction models (part 2): how to undertake an external validation study. BMJ. 2024;384: e074820. doi:10.1136/bmj-2023-074820

28. Riley RD, Snell KIE, Archer L, Ensor J, Debray TPA, van Calster B, et al. Evaluation of clinical prediction models (part 3): calculating the sample size required for an external validation study. BMJ. 2024;384: e074821. doi:10.1136/bmj-2023-074821

29. Vickers AJ, Elkin EB. Decision curve analysis: a novel method for evaluating prediction models. Med Decis Making. 2006;26: 565–574. doi:10.1177/0272989X06295361

